# "Exploring Cardiac Vector Propagation in Acute Myocardial Infarction: A Spatial Velocity Perspective"

**DOI:** 10.1101/2024.05.15.24307427

**Authors:** Tania Ghosal, Anjan Hembram, Imran Ahmed, Damodar Prasad Goswami, Anupam Bandyopadhyay, Arnab Sengupta

## Abstract

**Aims:** To objectively characterize the spatial-velocity dynamics of the QRS-loop in the vectorcardiogram (VCG) of patients with acute myocardial infarction (AMI).

**Methods:** VCG was constructed as space curve from three orthogonal ECG leads of 25 healthy individuals and 50 AMI patients. Spatial-velocity (SV) of the dynamic QRS loop was recorded, along with component attributes such as changing spatial distance (SD) and magnitude.

**Results:** Decreased SV and SD and altered spatial propagation patterns of ventricular vectors in AMI was recorded, with changes in specific axes based on infarct location. Reduced left-ventricular ejection-fraction indicates systolic dysfunction in AMI.

**Conclusion:** Altered spatial propagation patterns of vectors and ventricular dysfunction may be explained by complex interplay between mechanical and electrical factors in AMI.

**Declaration:**

a. The present manuscript has been submitted in the journal ***Future Cardiology***, and currently under peer review
b. None of the manuscript’s contents have yet been published.
c. All authors have read and approved the manuscript and approved its submission to MedRxiv, as preprint server. They also agreed the order of authorship.

## Introduction

Assessment of the coronary vascular anatomy and ventricular global and regional contractile performance constitutes the fundamental parameters for evaluating patients with Acute Myocardial Infarction (AMI). The clinical Electrocardiogram (ECG) is used for initial diagnosis and a limited prognostic assessment of the AMI [1,2].

AMI results in loss of structural and functional integrity of the different layers of heart, disruption of depolarization sources, macroscopic conduction anomaly, development of current of injury, altered excitability etc. [3]. Accordingly, a complex patho-physiological sequence ensues, that may progress towards electrical abnormalities, arrhythmia etc. Heart being a complex three-dimensional structure, there are numerous possible pathways along which the impulse can pass. A meticulous and thorough investigation of the cardiac electrical activities is essential as routine protocol in AMI. Accordingly, an in-depth examination of the instantaneous cardiac vectors propagated across the heart, becomes crucial.

The routine clinical ECG does not represent the instantaneous vectors generated at the heart and only shows the magnitude of the component of the main electrical vector along a particular lead axis. The magnitude, direction and the polarity of the instantaneous vectors cannot be known from the ECG. Thus, any small changes, early initial change or electrically compensated change of heart’s vector cannot be identified from the ECG [4]. Moreover, being scalar in nature, ECG does not explore the details of space domain, which is essential to understand the complex morphology of ventricular myocardium in health & disease.

Cardiac vectors are fundamentally time-varying current dipole propagating in the heart. Vectorcardiogram (VCG) represents a graphical representation of the trajectory of the tip of these vectors in three-dimensional space [4]. The VCG loops contain 3D recurring, near-periodic cardiac dynamics and can be projected onto different planes (X-Y, X-Z and Y-Z) to capture the time-space correlations in the two dimensions, or plotted as a static attractor in a 3D space that provides the topological relationships when all the variables are changing [5].

VCG is constructed by drawing the instantaneous vectors from a zero-reference point according to direction, magnitude, and polarity in the space. It is more informative and sensitive than conventional ECG because of its one extra degree of freedom as an evaluation tool of the physiology of cardiac dynamics. As a result, with the help of VCG it is possible to correlate even a minute electrophysiological alteration with the patho-physiological changes in diseases which is essential for early diagnosis [6]. There are limited number of studies of AMI evaluation using VCG as a tool [7-11]

In our earlier works [5,10], we assessed and characterized the static spatial morphology of the QRS loop with the help of three-dimensional vectorcardiography. We curiously found that, the spatial QRS loop lies in a planar orientation, which is lost in case of AMI. We attempted to offer some clarification of the emergence of planarity in normal and loss in the cases of AMI; however, detailed experimental and analytical studies are needed to explain the spatial morphologies of QRS loop and its changes in various conditions.

In the present study, we conducted a dynamic evaluation of ventricular electrophysiology using a high throughput Vectorcardiography (VCG) derived descriptor, i.e., Spatial Velocity of the Dynamic Vectorcardiogram loops, in patients suffering from acute myocardial infarction. Spatial characteristics of the electrocardiogram and the spatial electrocardiographic display on a continuous time basis is designed to uncover the dynamic detail of the temporo-spatial signals of the heart and has got great theoretical potential as a robust parametric expression of cardiac electrodynamics.

Despite the attractiveness of this mode of spatial data analysis and display, they are not yet widely adopted due to procedural complexity and a lack of standardization. With the advent of efficient computational and software tools, this issue has been mitigated.

Evaluation of Spatial Velocity (SV) of dynamic QRS loop in cases of AMI was done in order to understand the pattern of propagation of serial ventricular instantaneous vectors during the disease state. SV being a virtual velocity represented by the velocity of the moving cursor on the monitor screen, when the serial appearance of spatial points (x_1_-y_1_-z_1_, x_2_-y_2_-z_2_, ………x_n_-y_n_-z_n_) representing the tip of the instantaneous vectors, were animated. Electrophysiologically, SV signifies the rate of change of magnitude of the serial instantaneous vectors with respect to time & space [12]. It is essentially a mathematical construct, based upon certain information regarding the integrative effect of three orthogonal inputs [13].

An animated representation of the dynamic three-dimensional VCG loop is provided for visual display of the Spatial Velocity: https://youtu.be/gcA_8vQx8yc

An animated representation of the dynamic three-dimensional VCG loop of a normal volunteer is provided as supplementary material, for visual display of the Spatial Velocity.

Myocardial infarction is the severest form of coronary diseases causing a spectrum of cardiac anomalies as mentioned earlier, likely to affect the genesis and propagation of cardiac vectors. Depending upon the AMI subgroups (based on predominant site of injury, e.g., anterior wall, Inferior wall etc.) altered instantaneous vectors may follow a pattern of serial propagation which is likely to affect the SV. This altered instantaneous vector pattern may also lead to spatial shifting of the loop. All these things are likely to affect the SV.

However, although a few works were reported on the AMI evaluation by using SV [14-16], there is extensive scope of utilizing SV as an efficient tool of assessment of ventricular vector dynamics.

Component evaluation of the SV, with respect to three orthogonal axes namely SV_x_, SV_y_, SV_z_, give us detailed objective information about the spatial shifting of the VCG loop.

The present study is aimed at constructing the VCG by a simple technique as per Ray *et al* [10] to characterize temporo-spatial patterns of cardiac electrical activity and to objectively characterize the spatial velocity of the dynamic QRS loop of the VCG in normal individuals and patients of acute myocardial infarction. In this project, we attempted to capture the spatiotemporal characteristics of the VCG signals by viewing the animated cardiac vectors in real time on a computer screen instead of a static signal output. This method overcomes the drawbacks of conventional static VCG representation and provides simultaneous spatial and temporal resolutions.

We also propose and put forward certain possible mechanisms to explain the observed changes in vectorcardiographic spatial velocity in AMI.

## MATERIAL AND METHODS

We conducted a descriptive observational study on the extraction of features from the VCG in a group of patients with acute myocardial infarction, diagnosed at the Department of Cardiology, SSKM Hospital & Medical College Hospital, Calcutta, India. Cases were enlisted from the patients attended the emergency room diagnosed of Acute Myocardial Infarction, following clinical, electrocardiography and biochemical evaluation. However, patients associated with cardiovascular complications other than AMI which may affect the pattern of ECG like ventricular hypertrophy, conduction defect and other complication, or patients having non-cardiovascular diseases which may affect the ECG pattern of individual like hyperkalaemia, and patients with implanted gadgets (AICD, pacemaker etc.) were excluded from the study. A group normal subjects of devoid of any diagnosed disease condition, were included as control.

After receiving approval from the Institutional Ethics Committee of Institute of Postgraduate Medical Education & Research, Kolkata (Ref. No. IPGME&R/IEC/2021/114) and obtaining informed consent from each of the individual subjects, the study protocol was instituted. 25 healthy individuals aged between 18 and 65 years with a median age of 44 (64% male) and 50 patients with diagnosed cases of AMI aged between 39 and 76 (median age 59, 70% male) were recruited, and all necessary investigations including echocardiography were undertaken.

The study group with myocardial infarcts was divided into two major subgroups according to locations of the infarcts (anterior wall: AWMI, n = 24 and inferior wall: IWMI, n = 19). This classification was based on the conventional interpretation of the clinical electrocardiogram and findings of coronary angiogram. A few patients (n = 7) remain unclassified (non-STEMI, multiple wall extension etc.), and was not included in sub-group analysis.

The ECG signals were obtained by using a digital electrocardiograph (AD Instruments, Australia; sampled at 500 samples/s). Spatial vectorcardiograms were constructed using simultaneously recorded data of QRS complex from three orthogonal leads of I, aVF and V_2_ as detailed by Ray *et al* (10). The spatial velocity of the dynamic spatial QRS (SV_QRS_) of the Vectorcardiogram was recorded as the rate of change of amplitude of serial instantaneous ventricular vectors, expressed in numerical terms (mV/Sec). Throughout the research work, including clinical workup, data collection, recording, analysis etc., we strictly conformed to the World Medical Association Declaration of Helsinki regarding the ethical principles for medical research involving human subjects.

### Determination of Spatial Velocity of dynamic QRS loops (SV_QRS_)

The velocity of movement of the cursor point constituting the spatial vectorcardiographic QRS loop, which represents the gradient of change in magnitude of the consecutive instantaneous QRS vectors, with respect to time & space was estimated from the concurrently obtained QRS complex data from three orthogonal leads as stated above, using simple mathematical tool [5]. This is Essentially a scalar quantity, where directional changes are ignored in order to avoid negative magnitude. It is the point-to-point temporo-spatial gradient of the vectors of a representative 3D-QRS loop. Before summing up, the gradient values were transformed to their respective modulus values. The velocities along the three individual orthogonal axes were summed up to have the three-dimensional spatial velocity and the summation procedure constituted squaring, adding, and finally taking the positive square root. The squaring is necessary to make the entities independent of signs, summation for accumulating them and taking square root to keep the dimension balanced [12]. The final expression thus lands up to

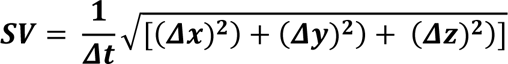

Where *Δx, Δy* and *Δz* are the change of the vector component in X, Y and Z axes respectively in the sampling interval *Δt*.

#### Evaluation of the Dynamic QRS Loop

The velocity components of SV_QRS_ in the X, Y and Z orientations, i.e. SV_X_ = *ΔX/Δt*, SV_Y_ = *ΔY/Δt* and SV_Z_ = *ΔZ/Δt* were also calculated.

The spatial magnitude of the instantaneous vector is calculated as,

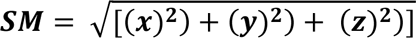

The spatial distance between the tips of the serial instantaneous vectors is calculated as,

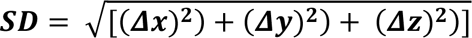

Recorded values the dynamic VCG parameter of SV and other attributes including SD, SM, ΔX, ΔY, and ΔZ and the echocardiographic parameter of Ejection fraction were analyzed using standard statistical tool. Comparison between control and total case groups and subgroups were done by unpaired t test. A p value of < 0.05 was taken as statistically significant.

3D-Spatial VCG of QRS loop and its planar projections on three orthogonal leads of a control individual and two illustrative cases of AWMI and IWMI were constructed for visual display of the ventricular vectors.

## RESULTS

The observed values of various SV parameters in the current project are in agreement with the data of the control volunteers, reported in the earlier papers (15, 16).

There is a noticeable decrease (12-25%) of the mean and maximum spatial velocity of the Dynamic QRS loop of VCG in most of the cases of AMI and its subgroups, as compared to controls (Table 1). However, there was no appreciable difference of the spatial velocities of 3D-QRS loops between the subgroups of AWMI and IWMI cases (Table 2).

**Table 1:**
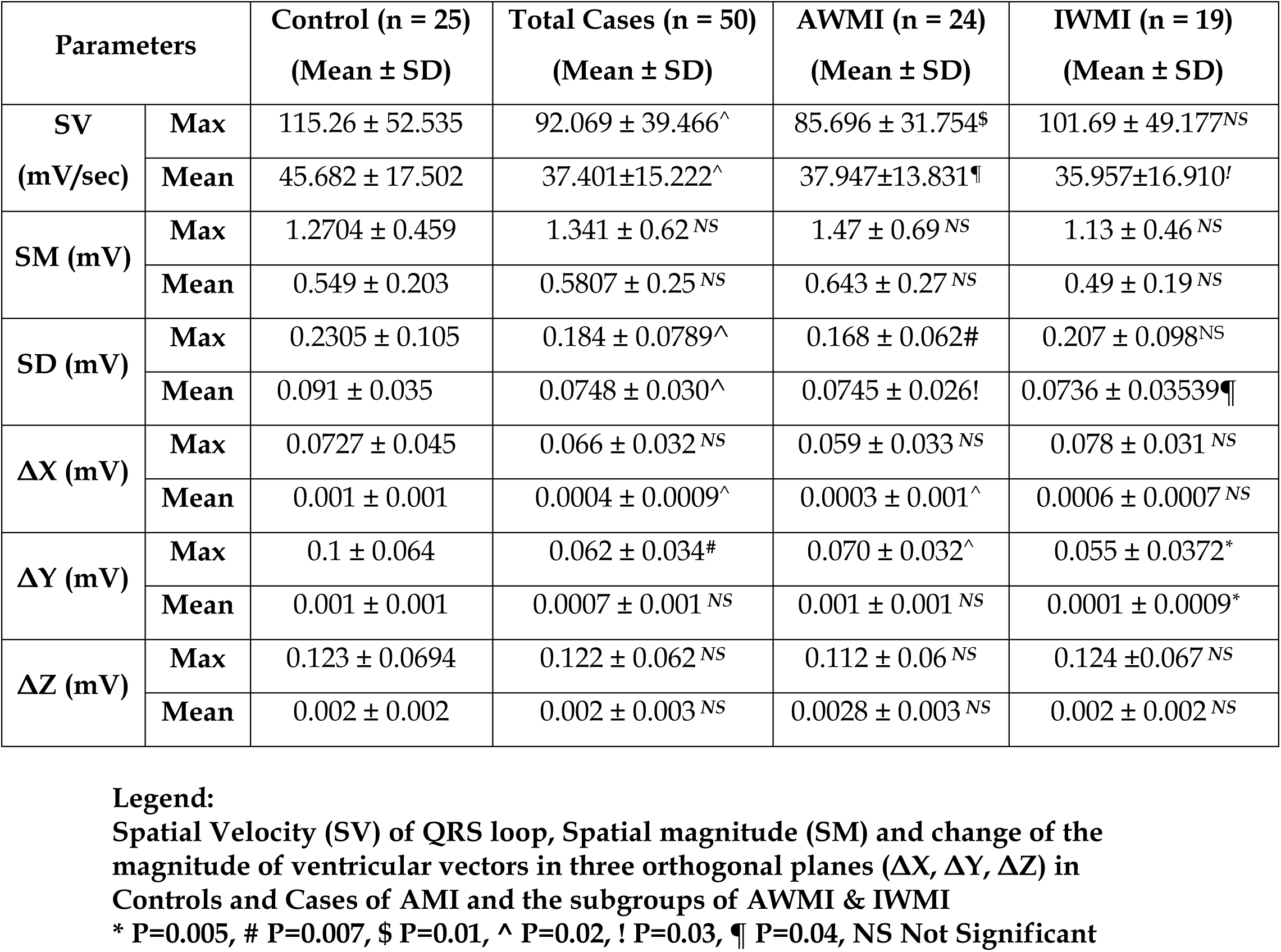
Comparison of Dynamic Vectorcardiographic Parameters between Control and Total Cases of AMI and AWMI & IWMI Subgroups.

**Table 2:** Comparison of Dynamic Vectorcardiographic Parameters between AWMI and IWMI Subgroups.

| Parameters | | AWMI (n = 24)<br>(Mean ± SD) | IWMI (n = 19)<br>(Mean ± SD) |
| --- | --- | --- | --- |
| SV<br>(mV/sec) | Max | 85.696 ± 31.754 | 101.69 ± 49.177 <sup>NS</sup> |
| | Mean | 37.947±13.831 | 35.957±16.910 <sup>NS</sup> |
| SD (mV) | Max | 0.168 ± 0.062 | 0.207 ± 0.098 <sup>NS</sup> |
| | Mean | 0.0745 ± 0.026 | 0.0736 ± 0.03539 <sup>NS</sup> |
| SM (mV) | Max | 1.47 ± 0.69 | 1.13 ± 0.46 <sup>NS</sup> |
| | Mean | 0.643 ± 0.27 | 0.49 ± 0.19 <sup>NS</sup> |
| ΔX (mV) | Max | 0.059 ± 0.033 | 0.078 ± 0.031<sup>\$</sup> |
| | Mean | 0.0003 ± 0.001 | 0.0006 ± 0.0007 <sup>NS</sup> |
| ΔY (mV) | Max | 0.070 ± 0.032 | 0.055 ± 0.0372 <sup>¶</sup> |
| | Mean | 0.001 ± 0.001 | 0.0001 ± 0.0009 <sup>@</sup> |
| ΔZ (mV) | Max | 0.112 ± 0.06 | 0.124 ±0.067 <sup>NS</sup> |
| | Mean | 0.0028 ± 0.003 | 0.002 ± 0.002 <sup>NS</sup> |

Spatial Velocity (SV) of QRS loop, Spatial magnitude (SM) and change of the magnitude of ventricular vectors in three orthogonal planes ( ΔX, ΔY, ΔZ) in the subgroups of AWMI and IWMI
@ P=0.006, \$ P=0.01, ¶ P=0.04, NS Not Significant

This general reduction of the spatial velocity in AMI cases must be due to the reduction of one or more velocity components of SV in the X, Y and Z orientations, i.e. SV_X_, SV_Y_ or SV_Z_. As the *Δt* component is fixed in this study (2 mSec, due to sample acquisition rate of 500 Samples/Sec.), the reduction in the spatial velocity is attributed to the decrease in the spatial distance (SD) between the tips of the ventricular instantaneous vectors due to reduction in the change of the amplitude of the serial instantaneous ventricular vectors (*ΔX, ΔY* and *ΔZ)* in various axial orientation (Table 1).

The spatial magnitudes (SM) of instantaneous ventricular vectors did not exhibit any statistically significant change in AMI patients as compared to controls.

It is evident from the Tables 1 and 2, that, the maximum change of the amplitude of the serial instantaneous ventricular vector in the Y axis (*ΔY_Max_*) was substantially reduced in AMI and its different subgroups e.g. AWMI and IWMI, as compred to controls. The mean change of the amplitude of the serial instantaneous ventricular vector in the Y axis (*ΔY_Mean_*) and X axis (*ΔX_Mean_*) were significantly reduced in IWMI and AWMI respectively. There was no change in *ΔZ* in AMI and its subgroups.

Comparison between subgroups (AWMI and IWMI), revealed that *ΔX_Max_* is substantially lower in AWMI than IWMI and both *ΔY_Max_* and *ΔY_Mean_* are markedly lower in IWMI than AWMI.

The evaluation of left ventricular Systolic function by assessment of the Left Ventricular Ejection Fraction (LVEF) was performed in AMI cases, which were compared with the standard normal Indian reference data, which is a published database of echocardiography of 773 normal Indian individuals [17].

There has been significant (P<0.0001) reduction of left ventricular ejection fraction in AMI cases (Table 3), as compared to Standard Indian Normal Reference (68.7 ± 4.8%) [17]. The reduction in LVEF is more (P<0.0001) in AWMI cases (37.7 ± 2.25%), than the IWMI cases (47 ± 6.06%).

**Table 3:** Comparison of Left Ventricular Ejection Fraction (%) between Standard Indian Normal Reference [17] and Cases of AMI and the subgroups of AWMI & IWMI.

| **Standard Indian Normal Reference [17]**<br>**(n = 773) (Mean ± SD)** | **Total Cases (n = 50)**<br>**(Mean ± SD)** | **AWMI (n = 24)**<br>**(Mean ± SD)** | **IWMI (n = 19)**<br>**(Mean ± SD)** |
| --- | --- | --- | --- |
| 68.7 ± 4.8 | 41.96 ± 5.94 | 37.7 ± 2.25 | 47 ± 6.06 |

The Left Ventricular Ejection Fraction (LVEF) of AMI and the sub-group of AWMi & IWMI was compared with the standard normal Indian reference data, which is a published database of echocardiography of 773 normal Indian individuals [17]. The difference between the normal and AMI & sub-groups were highly significant (P<0.0001). The reduction in LVEF is more in AWMi cases than the IWMI cases (P<0.0001).

The Figs. 1, 2 and 3 depict the 3D-QRS Spatial Vectorcardiogram and its planar projections in in three orthogonal planes (XY, YZ and XZ) in a control volunteer, and two illustrative cases of AWMI and IWMI respectively. The mutual distances between serial spatial points are apparently decreased in those illustrative cases.

**Fig. 1:**
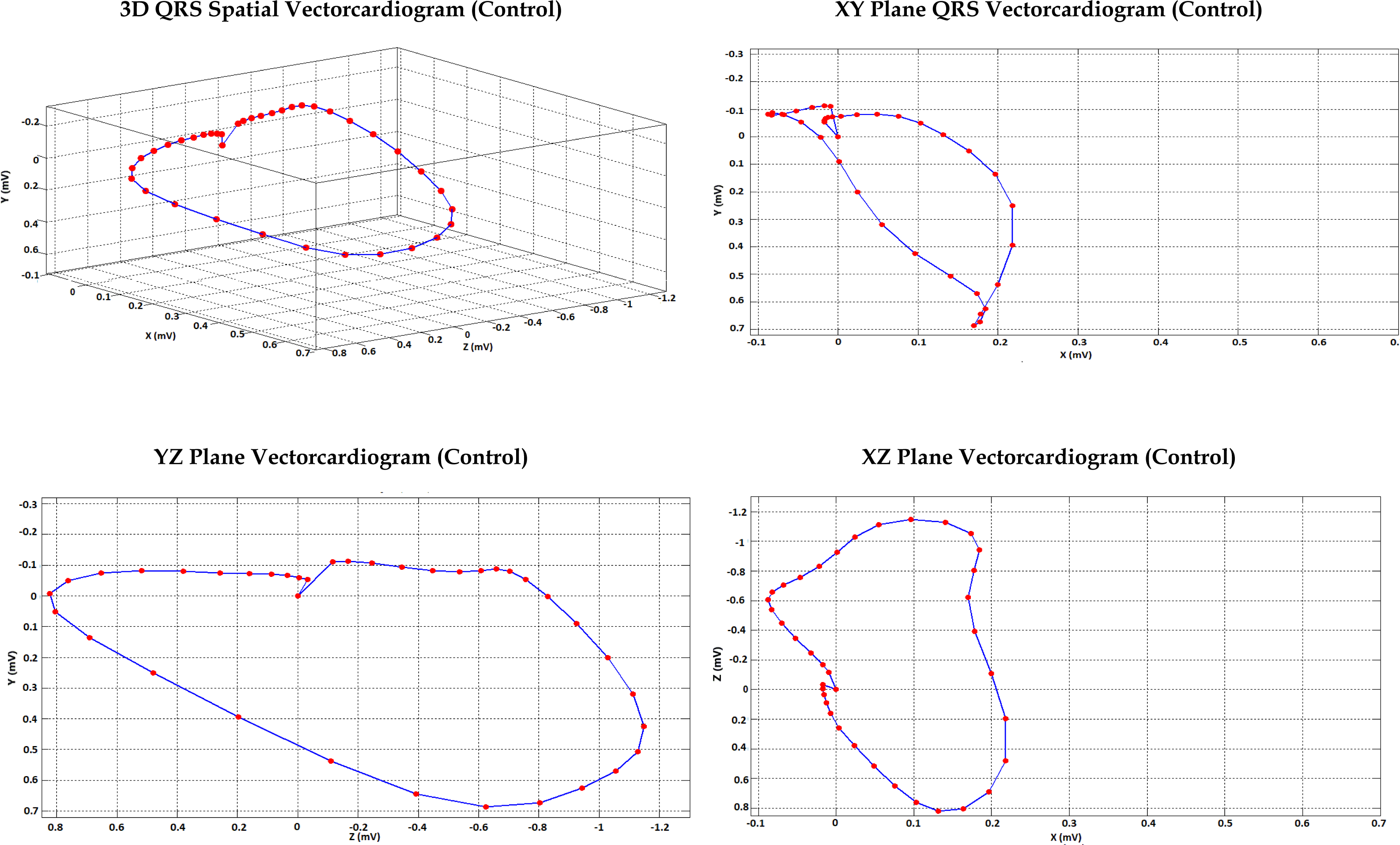
An Illustrative Vectorcardiogram in a Normal Control.

**Fig. 2:**
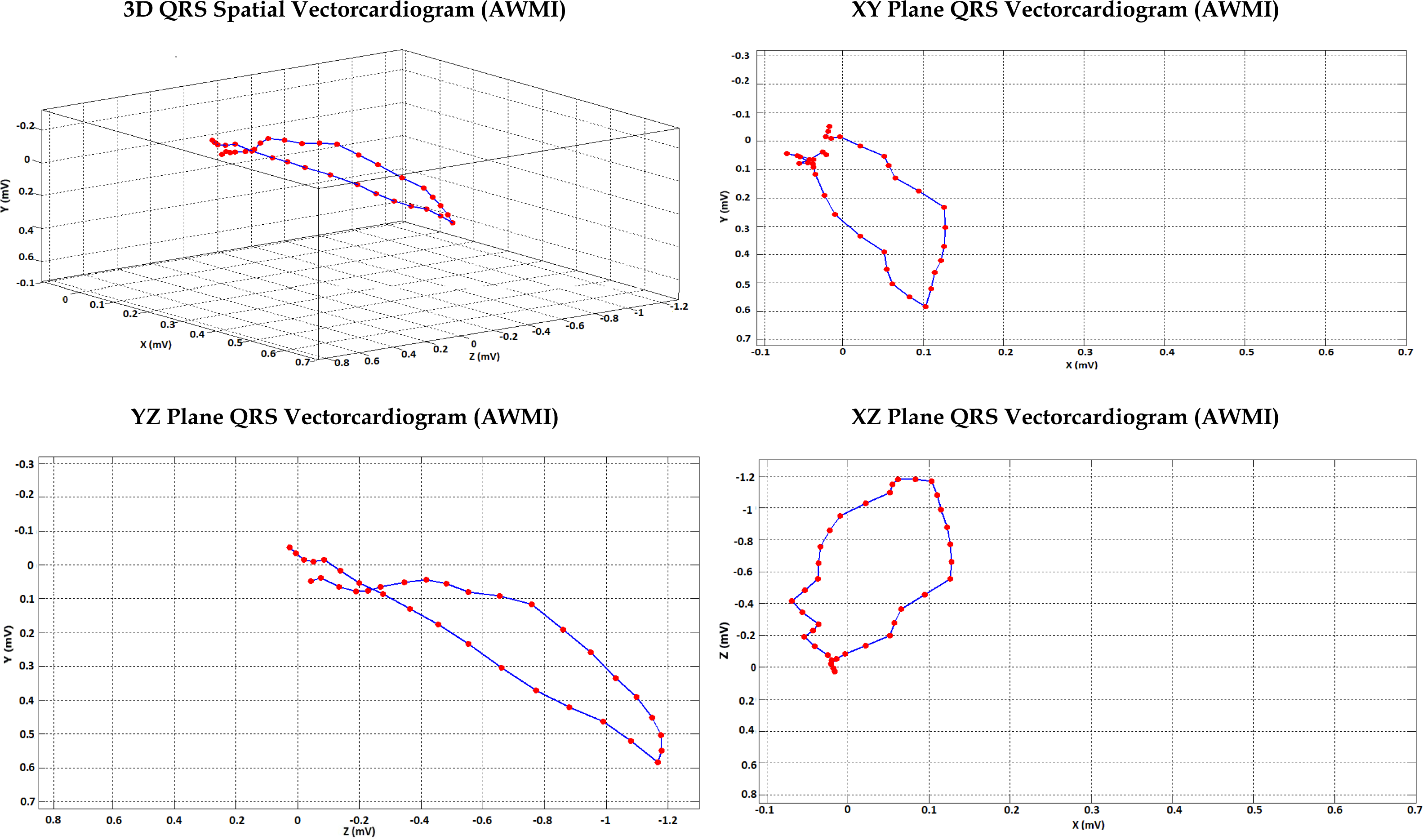
An Illustrative Vectorcardiogram in a Case of Acute Anterior Wall Infarction.

**Fig. 3:**
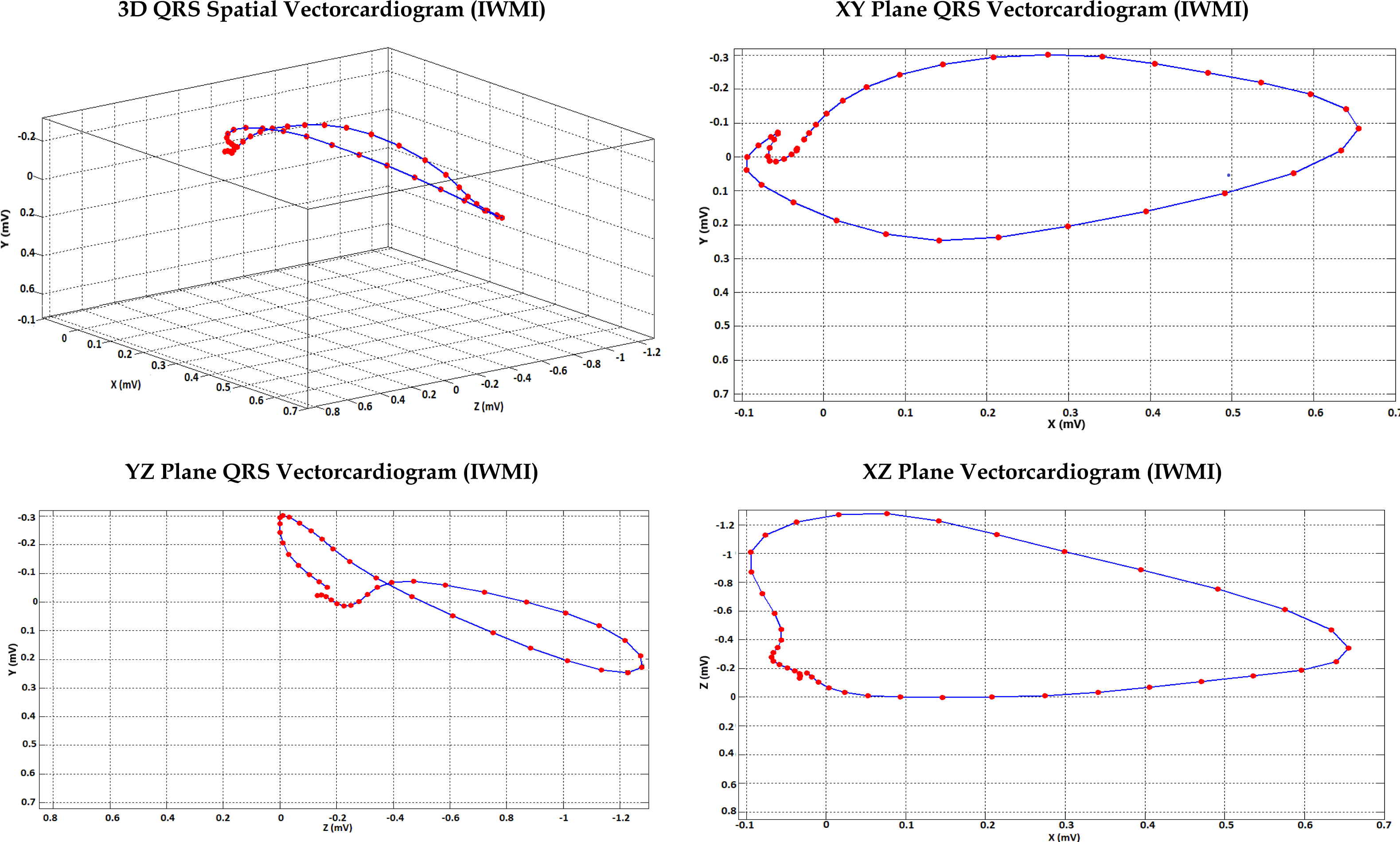
An Illustrative Vectorcardiogram in a Case of Acute Inferior Wall Infarction.

## Discussion

Dynamic vectorcardiography is the time-based curves of spatial parameters of the electrocardiogram that provides an optimal display of electrocardiographic data. This is apparently more informative than the conventional and static vectorcardiography loop.

Elucidation of spatial magnitude, recording of spatial distance, calculation of changing magnitude of serial spatial vectors, spatial velocity, and orientation of the vectors provide crucial insight into the dynamics of cardiac electrophysiology.

Exploration of the characteristics of the serial vectors in space in AMI cases and the control volunteers yielded interesting observations.

The overall findings in the present study indicate a general reduction in the spatial velocity of the Dynamic QRS loop of VCG in most of the cases of AMI and its subgroups, as compared to controls, without any change of the spatial magnitudes of instantaneous ventricular vectors (Table 1, 2). This decrease in the spatial velocity is due to reduced spatial distance between the tips of the instantaneous vectors in cases of AMI leading to decrease in the change of the amplitude (*ΔX, ΔY, ΔZ*) of the serial instantaneous ventricular vectors in cases of AMI (Figs. 1, 2 and 3). However, all those decreases were not always attended the level of statistical significance, at the current degree of freedom in this limited sample size of the study. Specifically, the significant vector changes were found in the Y-Axis in IWMI (*ΔY*) and X-Axis in cases of AWMI (*ΔX*), as compared to controls. No significant changes were observed in the Z-Axis. Accordingly, decrease in the change in the vector magnitudes were seen in superior-inferior orientation in cases of IWMI and side-to-side orientation in AWMI, and no significant anterior-posterior changes of serial ventricular vectors were observed in cases of myocardial infarction. Without any change in the spatial magnitude (SM) (Table 2), reduced SD as well as decrease in *ΔY* and *ΔX* in IWMI and AWMI respectively seem to arise from the fact of greater approximation and close clustering of the instantaneous ventricular vectors in AMI cases, as compared to controls.

Also there has been significant reduction in left ventricular ejection fraction due to compromised pumping ability of heart in AMI cases, as compared to Standard Indian Normal Reference [17] (68.7 ± 4.8%). The reduction in LVEF is greater in AWMI cases (37.7 ± 2.25%), than the IWMI cases (47 ± 6.06%).

There is substantial concordance of the findings with the existing literature [18, 19], and as the predominant involvement of left ventricle myocardium takes place in AWMI, the systolic function is more compromised in this subgroup.

This systolic ventricular dysfunction pattern is a characteristic feature of AMI. The decrease in LVEF in AMI and the reduced change of the amplitude of the serial instantaneous ventricular vectors in cases of AMI has got crucial *effect & cause* perspective.

In the present study, a 20% to 30% reduction in Left Ventricular Ejection Fraction (LVEF) in AMI causes increased retention of blood in ventricle and substantial rise in end-diastolic ventricular volume (EDVV) or preload, resulting in passive stretch and an increased wall stress in circumferential orientation along the fiber direction. This increased wall stress due to abnormal loading acting on the muscle comprising the ventricular wall, might lead to ventricular electrical alterations.

In order to explain and justify the phenomenon of close approximation and clustering of the tips of the instantaneous ventricular vectors and systolic ventricular dysfunction in AMI cases, we would like to put forward certain basic physiological aspects of ventricular electrical & mechanical properties.

The feature of systolic dysfunction in acute myocardial infarction is essentially due to the loss of structural and functional integrity of the different layers of heart [3]. There is alteration of the magnitude of the action potentials leads to a change of the intensity of the line density of the membrane current [3]. Consequently, the increased end systolic volume (ESV), due to reduced ejection fraction causes increase in preload (EDVV) of the ventricle. The increase preload increases the ventricular volume & ventricular wall stress, which is directly proportional to the diameter of the ventricle and the ventricular pressure. Increased ventricular wall stress is responsible for the adverse remodeling process [20].

The heart is continuously subjected to various mechanical forces. Increased stress induces the recruitment of ion channels from intracellular storage pools to the plasma membrane [21]. The incorporation of channels into the membrane causes changes in the electrical activity of the myocyte and may be an important way for cells to adapt to increased mechanical forces.

It is known that the heart depends on electrical depolarization to trigger mechanical contraction, and mechanical perturbations have also long been recognized to affect cardiac electrical activities. [20]. The process by which myocyte electrical activation leads to mechanical contraction is known as excitation-contraction coupling (ECC), while the process by which a mechanical alteration influences cardiac electrical activity is referred to as mechano-electric feedback (MEF). Both phenomena are manifested at cellular and whole heart scales.

Ventricular electrical alteration is frequently seen in patients with ventricular dysfunction, ventricular volume, or pressure overload, or dyssynergistic ventricular contraction and relaxation [20-22], which are the examples of MEF.

This *Cardiac Electro-Mechanical Coupling* is the electrical and mechanical systems, those are intimately linked by the anisotropic microstructure of the heart, that regulates the spread of the electrical wave and the resulting deformation. Each system also regulates the other via a complex web of feedback mechanisms, with the extent of deformation dependent on the frequency of electrical activation and the electrical properties of the cell dependent on deformation [22].

In AMI, there is approximation and close clustering of the instantaneous ventricular vectors, due to decreased spatial distance as a result reduced rate of change of amplitude of serial instantaneous ventricular vectors.

The increase in preload, i.e., raised diastolic filling pressure moves the relationship to the right on the stroke volume-preload curve, like the effect of volume overload. The increase in the chamber dimension increases wall stress and along with associated tachycardia in AMI, the ventricles develop higher myocardial oxygen demand and relative ischemia. Ischemia affects cellular depolarization by altering chemical gradients and membrane conductance to ions. These changes alter the propagation pattern of ventricular vectors in AMI. Thus, in abnormal loading of the heart, the stress-strain relationship changes, an increased wall stress and associated ischemia in AMI potentially alter the pattern of propagation of the spatial vectors.

Ventricular stress-strain relationships during cardiac cycle affect the electrical activities as a part of MEF. Mechanical loading promotes meandering through MEF. Several ion channels are regulated by mechanical forces, which directly affect the gating of the channel [23] or indirectly activate intracellular signaling pathways to alter channel properties [24]. The response to stretch has been well documented that includes, activation of stretch activated ion channels [21], and Stretch activated Receptors [25].

Study of spatial vectors by simple mathematical operation of the conventional ECG data, gives valuable insight into the pathophysiology AMI progression. A real-time display of dynamic VCG loop on the monitor along with point-to-point determination of various parameters like SV, SM, SV_X_, SV_Y_, SV_Z_ and *ΔX, ΔY & ΔZ* etc. will provide valuable insight.

Regular evaluation and follow ups of the serial spatial ventricular vectors in AMI shall yield valuable information to understand the progress and prognosis of the disease process. The current method is simple, easy, and inexpensive, and a novel method to study the ventricular stress and MEF of the cardiac system.

## Limitations

There was limited sample size of the current study. This was due to ethical consideration related to the management priority of AMI patients. Also, echocardiographic data of normal controls were collected from a published database of normal Indian individuals.

A detailed study with greater sample size of each subgroup population is needed to assess the progress pattern of the spatial vectors.

## Conclusions

We conducted a dynamic evaluation of ventricular electrophysiology by recording the Spatial Velocity of the Vectorcardiogram loops, in patients suffering from acute myocardial infarction. There was significant reduction of Spatial Velocity in cases of AMI, due to decreased spatial distance and reduced rate of change of magnitude of the serial ventricular vectors. Accordingly, there is greater approximation and close clustering of the instantaneous ventricular vectors in AMI cases, as compared to controls. There is also reduced ejection fraction indicating a systolic dysfunction in AMI, which leads to an increase in end-systolic volume resulting in elevated preload and chamber dimension of the ventricle along with increased ventricular wall-stress. With associated tachycardia in AMI, the ventricles develop higher myocardial oxygen demand and relative ischemia. These changes along with the mechano-electric feedback exerted by the ventricular wall-stress, collectively affect the spatial propagation pattern of ventricular vectors in AMI with close clustering of the ventricular vectors and reduction of the spatial velocity.

## Supporting information

Supplemental Video showing Spatial Velocity of QRS Loop

## Data Availability

All the data produced in the present study are available upon reasonable request to the authors

https://youtu.be/gcA_8vQx8yc

## Sources of support in the form of grants

Science & Engineering Research Board (SERB) and Dept. of Science & Technology Grant Ref. No. CRG/2020/004318, Govt. of India.

## Declaration of Competing Interest

None.

## Supplementary media

On-line video link https://youtu.be/gcA_8vQx8yc

## Notes

**Conflict of interest disclosure** Authors have no conflict of interest. The present research is supported in the form of grants of the Science & Engineering Research Board (SERB), Dept. of Science & Technology, Govt. of India. Grant Ref. No.: CRG/2020/004318/BHS. There are no financial conflicts of interest to disclose.

### Competing Interest Statement

The authors have declared no competing interest.

### Funding Statement

The present research is supported in the form of grants of the Science & Engineering Research Board (SERB), Dept. of Science & Technology, Govt. of India. Grant Ref. No.: CRG/2020/004318/BHS. There are no financial conflicts of interest to disclose.

### Author Declarations

The ethical approval for studies involving human subjects was obtained from the Institutional Ethics Committee of the Institute of Postgraduate Medical Education & Research, Calcutta, India, vide No. IPGME&R/IEC/2021/114 dated 06/02/2021.

